# Calcium channel blocker amlodipine besylate is associated with reduced case fatality rate of COVID-19 patients with hypertension

**DOI:** 10.1101/2020.04.08.20047134

**Authors:** Lei-Ke Zhang, Yuan Sun, Haolong Zeng, Yudong Peng, Xiaming Jiang, Wei-Juan Shang, Yan Wu, Shufen Li, Yu-Lan Zhang, Liu Yang, Hongbo Chen, Runming Jin, Wei Liu, Hao Li, Ke Peng, Gengfu Xiao

## Abstract

The coronavirus disease (COVID-19) caused by the novel severe acute respiratory syndrome coronavirus 2 (SARS-CoV-2) has now spread to more than 100 countries posing as a serious threat to the public health on a global scale. Patients with comorbidity such as hypertension suffer more severe infection with elevated case fatality rate. Development of effective anti-viral drug is in urgent need to treat COVID-19 patients. Here we report that calcium channel blockers (CCBs), a type of anti-hypertension drugs that are widely used in the clinics, can significantly inhibit the post-entry replication events of SARS-CoV-2 in vitro. Comparison with two other major types of anti-hypertension drugs, the angiotensin converting enzyme inhibitors (ACEI) and angiotensin II receptor blockers (ARB), showed that only CCBs display significant anti-SARS-CoV-2 efficacy. Combined treatment with chloroquine and CCBs significantly enhanced the anti-SARS-CoV-2 efficacy. Retrospective clinical investigation of COVID-19 patients revealed that the CCB amlodipine besylate administration was associated with reduced case fatality rate of patients with hypertension. Results from this study suggest that CCB administration for COVID-19 patients with hypertension as the comorbidity might improve the disease outcome.

## Introduction

The severe acute respiratory syndrome coronavirus 2 (SARS-CoV-2) is the causative pathogen of the novel coronavirus disease (COVID-19) that recently occurred in Wuhan, China in late December 2019 (*1, 2*). SARS-CoV-2 infection induced similar symptoms with SARS-CoV including fever, cough, dyspnea, etc and can result in multiple organ dysfunction syndrome and death in severe cases (*3*). The virus has caused a global pandemic transmission posing as a serious threat to the public health. As of Mar 19, 2020, there are over 203,000 confirmed COVID-19 cases with more than 8,100 deaths from SARS-CoV-2 infection around the world. Due to its high transmissibility and severe infection outcome, the World Health Organization has declared the SARS-CoV-2 a public health emergency of international concern. The rapid transmission of SARS-CoV-2 raises the concern whether it will become a seasonal coronavirus like hCoV-229E, OC43, NL63, and HKU1, however with a much higher mortality rate. Development of effective anti-viral drugs is urgently needed to contain the current transmission of SARS-CoV-2 and to counteract its potential re-emergence in the future.

So far, no antiviral drug for SARS-CoV-2 has been officially proved to be effective in treating COVID-19 patients. Compared with de-novo drug development, which normally takes years of development and evaluation, repurposing preexisting drugs that are in clinical use to treat virus infection is one of the most effective strategies for developing drug against emerging viruses (*4*). Our recent study has reported that remdesivir, favipiravir and chloroquine (CQ) have distinct anti-SARS-CoV-2 effect in vitro (*5*). Remdesivir was first developed for treating Ebola virus (*6*), and showed strong anti-SARS-CoV and MERS-CoV activity in vitro (*7*)and in mouse model (*8*). A randomized controlled trial has been initiated to assess the efficacy and safety of remdesivir to treat COVID-19 and the result is expected to be released in April (*4*). Favipiravir is an approved anti-influenza drug for clinical use in Japan and very recently in China. Similar with remdesivir, favipiravir has also being registered in clinical trial to evaluate its efficacy in treating COVID-19(*4*). CQ is an anti-malaria drug that has been developed in the 1940’s with a safe record in clinical administration (*9*). Given its approved status it was quickly tested in clinics and a recent study reported its potential benefits in treating COVID-19 patients (*10*). These progresses strongly support the endeavor of repurposing approved drugs for COVID-19 treatment.

The most affected COVID-19 patients are the elderly who often have comorbidities such as hypertension, diabetes, cardiovascular disease, etc (*11*). These patients suffer more severe infection outcome with significantly higher case fatality rate (*11*). The current therapeutic regime is largely symptomatic treatment and specific evaluation of drug treatment for COVID-19 patients with different comorbidities is still lacking. Identification of more drug candidates with anti-SARS-CoV-2 efficacy would help to provide more options from which safe and effective drugs can be selected and/or combined for personalized medication for the patients on an individual level.

Calcium channel blockers (CCBs) are widely used in the clinics for treating hypertension, angina pectoris, supraventricular arrhythmias (*12*). Recently, CCBs were also reported to have anti-viral effect against several emerging viruses including bunyaviruses, arenaviruses and flaviviruses (*13-15*). About 30% of SARS-CoV-2 patients have hypertension as comorbidity and these patients suffer the case fatality rate of up to 14% (*11, 16*) urging that effective drug treatment for these patients needs to be evaluated. Recently, a concern was raised about whether administration of anti-hypertension drugs of ARB or ACEI to COVID-19 patients would worsen the disease progression through up-regulation of ACE2 expression level and result in more severe SARS-CoV-2 infection (22). In this study we tested a panel of anti-hypertension drugs that are in clinical use and found that the CCBs benidipine HCI and amlodipine besylate have significant anti-viral effect in vitro. Retrospective clinical investigation showed that amlodipine besylate was associated with reduced case fatality rate of COVID-19 patients with hypertension. These results provide valuable reference for selecting drug treatment for COVID-19 patients with hypertension as the underlying comorbidity.

## Results

### CCBs inhibit SARS-CoV-2 infection in vitro

To test whether CCBs can inhibit SARS-CoV-2 replication, Vero E6 cells were treated with a panel of 9 clinically approved CCBs, and then infected with SARS-CoV-2 at a multiplicity of infection (MOI) of 0.05. At 24 hours post infection (p.i.), copy numbers of viral RNA in the supernatant were measured with qRT-PCR (Figure 1A), and the intracellular level of virus infection was monitored by immunofluorescence with an antibody against virus NP protein (Figure 1B). We found that four CCBs, benidipine HCI, amlodipine besylate, cilnidipine and nicardipine HCI, significantly inhibited SARS-CoV-2 replication (Figure 1). Experiments with serial concentrations of drug treatment revealed that these four CCBs inhibited SARS-CoV-2 replication in a dose-dependent manner, without causing strong cytotoxic effect (Figure 2A). The half maximal inhibitory concentrations (IC_50_) of benidipine HCI, amlodipine besylate, cilnidipine, nicardipine HCI were 3.81, 4.17, 11.58 and 13.32 μM, respectively, and the half cytotoxic concentration (CC_50_) of all four drugs were calculated to be above 100 μM. The drug selection index (SI) of these four CCBs was calculated to be > 26.25, > 23.98, >8.64 and >7.51, respectively (Figure 2A). Similar inhibition effects of these four CCBs were also observed on the human hepatocyte cell line Huh7 (Supplementary Figure S1).

**Figure 1.**
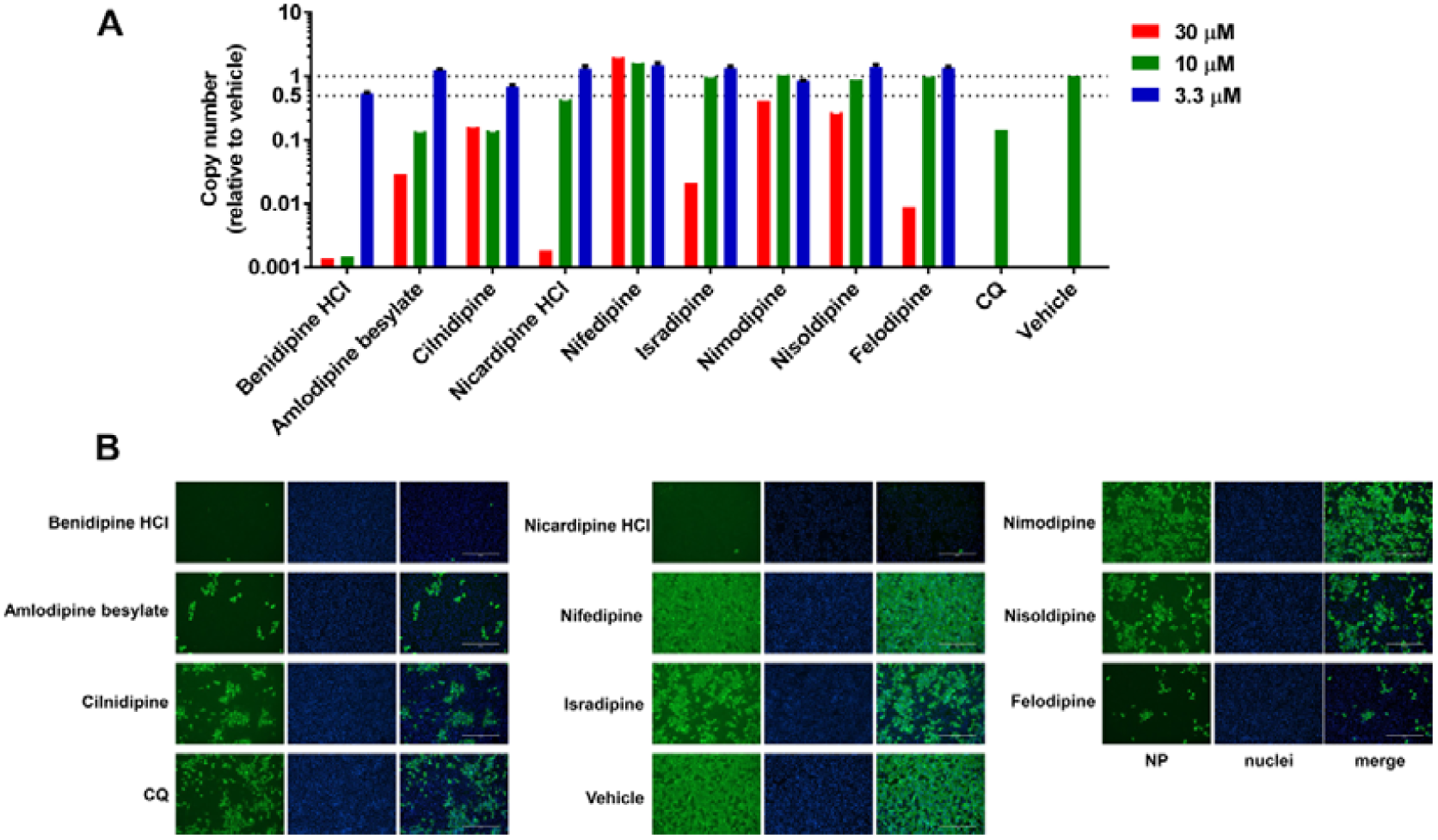
Evaluation of anti-SARS-CoV-2 activity of a panel of CCBs. Vero E6 cells were treated with indicated concentrations of compounds and infected with SARS-CoV-2 at an MOI of 0.05, and at 24 hours p.i., supernatant was collected and cells were fixed. Chloroquine (CQ, 5 μM) was used as positive control. (**A**) Viral copy number in the supernatant was measured with quantitative RT-PCR; (**B**) intracellular NP level in cells treated with 30 μM indicated compound was monitored with immunofluorescence. The experiments were done in triplicates, and data shown are means ± SD. Bars: 400 μm.

**Figure 2.**
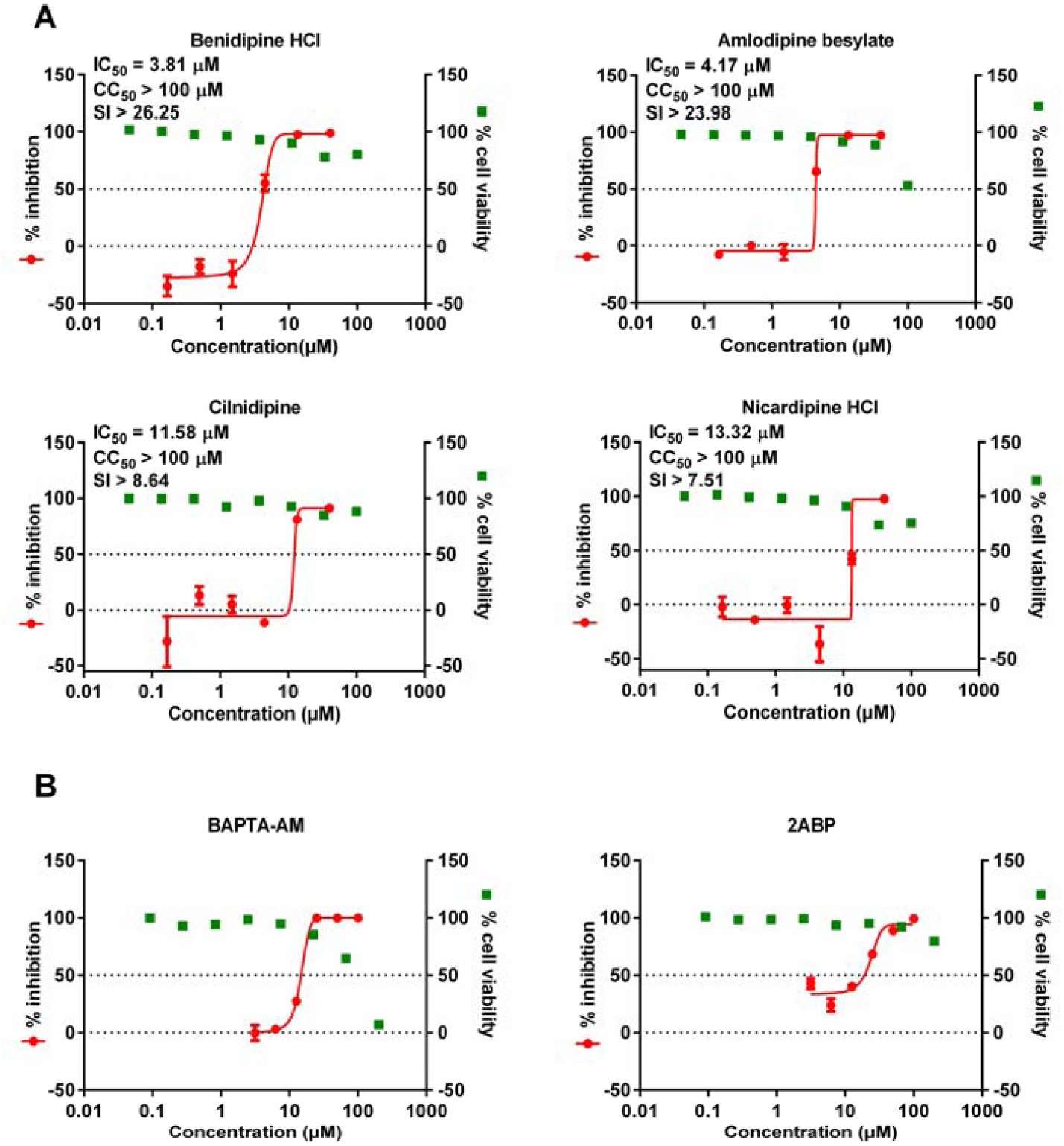
Dose dependent effects of benidipine HCI, amlodipine besylate, cilnidipine, nicardipine HCl, BAPTA-AM and 2ABP on SARS-CoV-2 replication. Vero E6 cells were treated with indicated concentrations of compounds and infected with SARS-CoV-2 at an MOI of 0.05, and at 24 hours p.i., supernatant was collected and viral copy number in the supernatant was measured with quantitative RT-PCR. Cell viability was measured with CCK8 assay. The left Y-axis of the graph indicates mean % inhibition of virus, while right Y-axis represents mean % cell viability. The experiments were done in triplicates, and data shown are means ± SD. The IC_50_ and CC_50_ values were calculated by Graphpad Prism 6.0.

Since CCBs block intracellular calcium influx, we analyzed whether the anti-SARS-CoV-2 effect of CCBs is related with reduced intracellular calcium level. Intracellular calcium level can be reduced through treatment with calcium chelator 1,2-Bis(2-aminophenoxy)ethane-N,N,N’,N’-tetraacetic acid tetrakis(acetoxymethyl ester) (BAPTA-AM) or 2-Aminoethyl Diphenylborinate (2APB), a membrane permeable blocker of the inositol 1,4,5-trisphosphate (IP3)-induced Ca^2+^ release (*17*). Vero E6 cells were treated with serial concentrations of BAPTA-AM or 2APB, and then infected with SARS-CoV-2. At 24 hours p.i., copy numbers of viral RNA in the supernatant were measured with qRT-PCR. As shown in figure 2B, addition of BAPTA-AM or 2APB also significantly inhibited virus replication in a concentration dependent manner, confirming the dependence role of intracellular Ca^2+^ for SARS-CoV-2 replication.

### CCB inhibits viral replication at the post-entry stage

To define the event of virus infection that was inhibited by CCBs, time-of-addition assay of drug treatment was performed. The CCB benidipine HCI was chosen for further analysis as it has the lowest effective concentration and the highest SI index of the 4 tested CCBs. Benidipine HCI or 2ABP were added during virus entry, 2-hours post virus infection or through-out virus infection (Figure 3A). The virus production in the supernatant was measured with qRT-PCR and the intracellular NP expression level was determined with western blot and immunofluorescence analysis with the NP antibody. As shown in figure 3B-D, addition of drug through-out virus infection or 2-hours after virus entry strongly inhibited virus production, while addition of drug during virus entry did not inhibit virus replication. Notably, compared with drug treatment throughout virus infection, addition of drugs 2-hours after virus entry had slightly lower inhibition efficacy (Figure 3 B,C). Whether this is due to rapid onset of virus replication within the first 2-hours that was not fully blocked by following drug treatment still needs further characterization. Nevertheless, these results indicate that benidipine HCI and 2ABP mainly inhibit virus infection at a stage after virus entry, potentially during virus genome replication/transcription.

**Figure 3.**
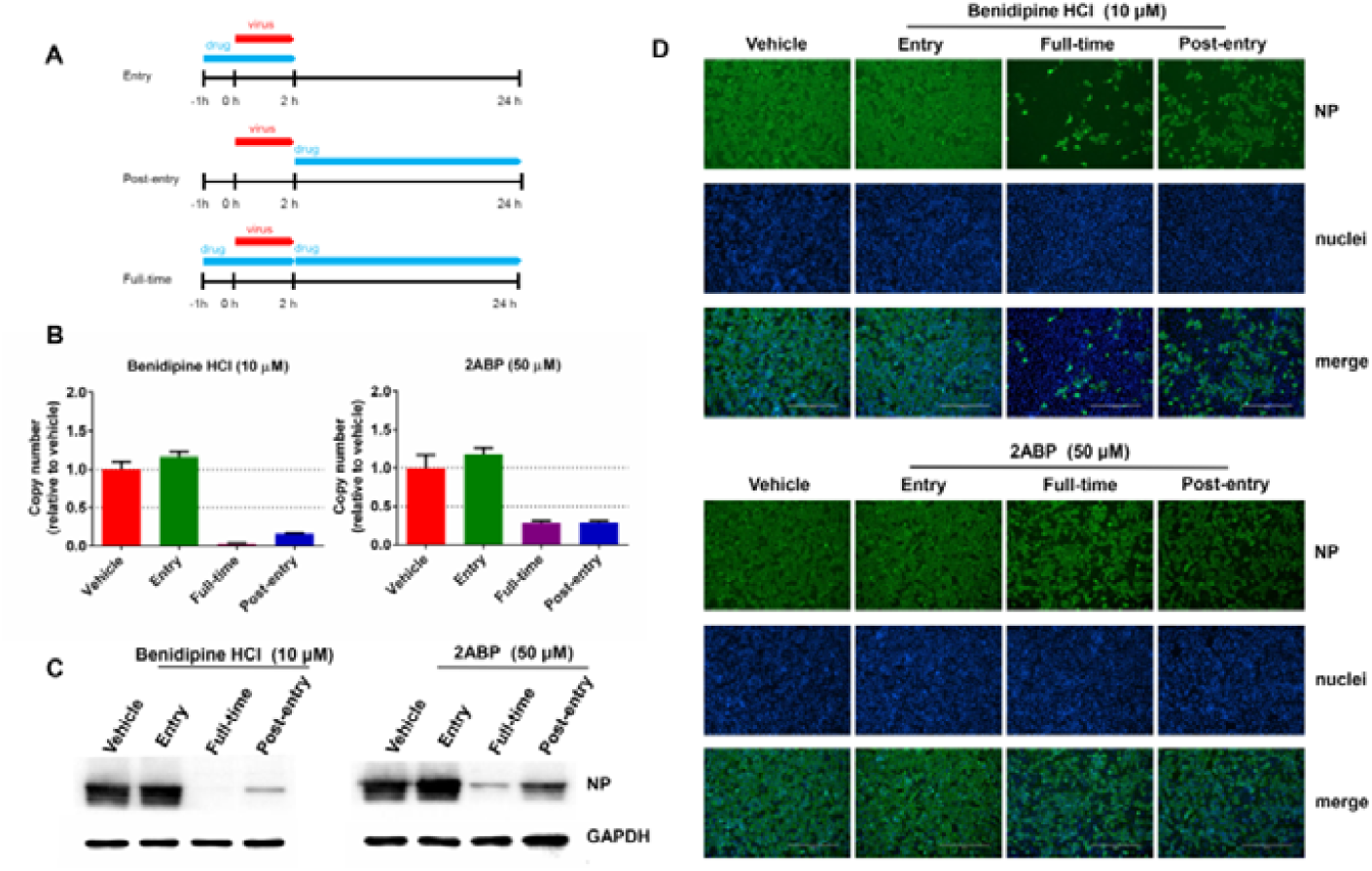
Time-of-addition experiment of benidipine HCI and 2ABP. (**A**) For “Full-time” treatment, Vero E6 cells were pre-treated with compounds for 1 hour, and then infected with virus. At 2 hours p.i., the supernatant was removed, and the cells were cultured with compound-containing medium until the end of the experiment. For “Entry” treatment, Vero E6 cells were pre-treated with compounds for 1 hour, and then infected with virus. At 2 hours p.i., the supernatant was removed, and the cells were cultured with fresh culture medium until the end of the experiment. For “Post-entry” experiment, Vero E6 cells were infected with virus, and at 2 hours p.i., cells were treated with compound-containing medium until the end of the experiment. For all these experiments, Vero E6 cells were infected with SARS-CoV-2 at an MOI of 0.05, and virus copy number in the supernatant was quantified by quantitative RT-PCR (**B**) and NP expression in infected cells was analyzed by western blot (**C**) and immunofluorescence with NP antibody (**D**) at 24 hours p.i.. The Y-axis of the graph represents mean % inhibition of virus. The experiments were performed in triplicates.

### CCBs but not ARBs or ACEIs display inhibitory effect against SARS-CoV-2 replication

Angiotensin II receptor blockers (ARB), angiotensin converting enzyme inhibitors (ACEI) and CCBs represent three major types of anti-hypertension drugs that are in clinical use (*18*). We next analyzed whether the ARB and ACEI anti-hypertension drugs can also inhibit SARS-CoV-2 replication. Representative ARBs (losartan potassium, valsartan) or ACEIs (enalaprilat dihydrate, enalapril maleate) that are widely used in the clinics (*18*) were chosen for the evaluation of potential anti-viral effect. Vero E6 cells were treated with serial concentrations of drug compounds and infected with SARS-CoV-2 at an MOI of 0.05. At 24 hours p.i., viral copy number in the supernatant was measured with qRT-PCR and cell viability was measured with CCK-8 assay. As shown in figure 4, in contrast to the distinct inhibition efficacy against SARS-CoV-2 of CCBs, the selected ARBs or ACEIs did not show any significant inhibition effect. These results suggested that of the three types of anti-hypertension drugs only CCBs have significant anti-SARS-CoV-2 efficacy.

**Figure 4.**
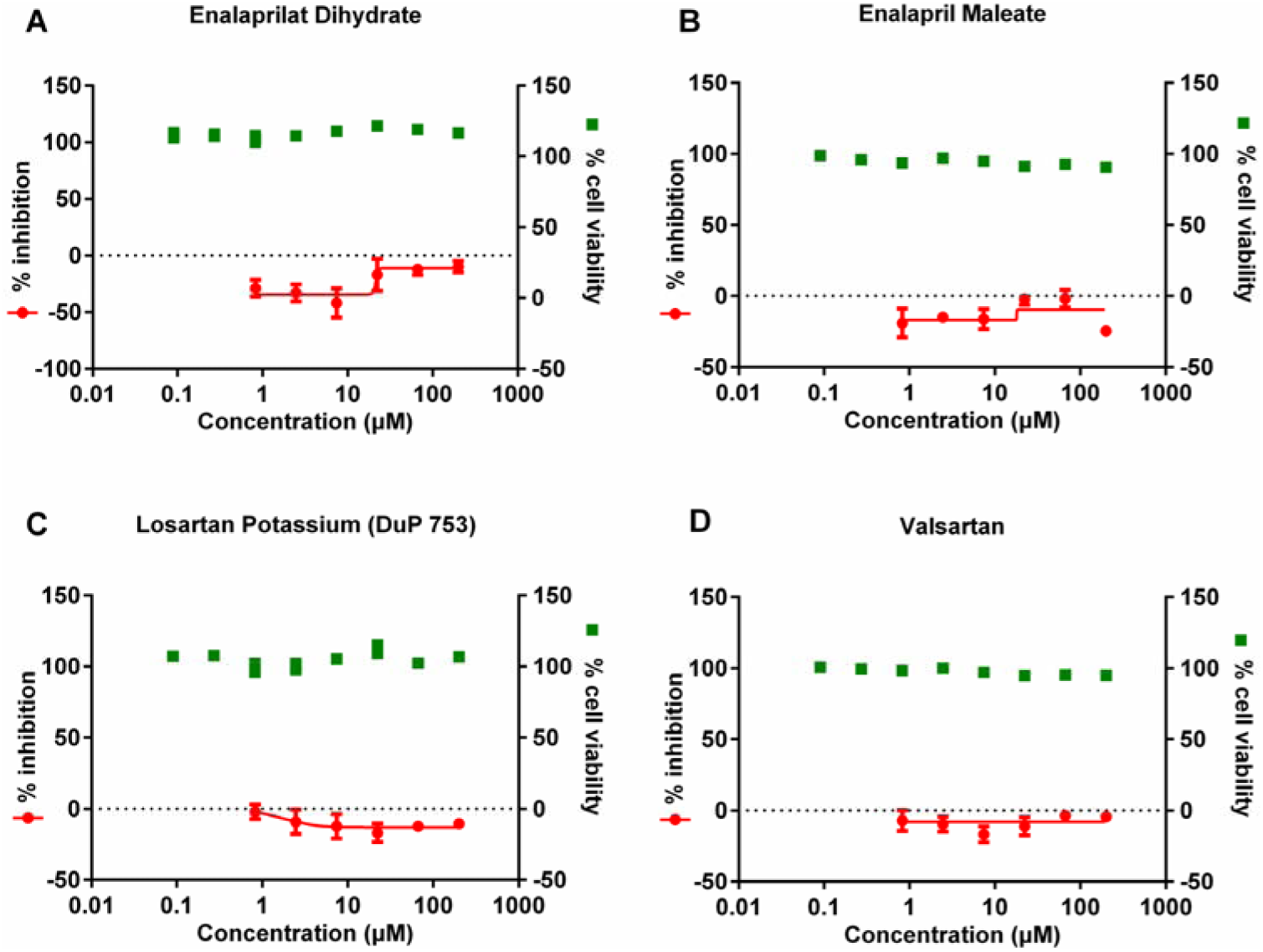
Effect of drug treatment with two ACEIs (enalaprilat dihydrate, enalapril maleate) or two ARBs (losartan potassium, valsartan) on SARS-CoV-2 replication in vitro. Vero E6 cells were treated with indicated concentrations of compounds and infected with SARS-CoV-2 at an MOI of 0.05. At 24 hours p.i., supernatant was collected and viral copy number in the supernatant was measured with quantitative RT-PCR. Cell viability was measured with CCK8 assay. The left Y-axis of the graph indicates mean % inhibition of virus, while right Y-axis represents mean % cell viability. The experiments were performed in triplicates, and data shown are means ± SD.

### Combined application of chloroquine (CQ) with CCB resulted in enhanced anti-SARS-CoV-2 effect

CQ was recently reported to inhibit the entry stage of SARS-CoV-2 replication (*5*). Considering that CCB may inhibit SARS-CoV-2 at the post-entry stage, we analyzed whether the combined application of CQ and CCB would lead to a more distinct inhibition effect. CQ and CCB were added separately or in combination to the Vero E6 cells followed by virus infection with SARS-CoV-2 at the MOI of 0.05. At 24 hours p.i., copy numbers of viral RNA in the supernatant were measured with qRT-PCR and the intracellular level of virus infection was monitored by immunofluorescence with the NP antibody. As shown in figure 5, while separate application of CQ or benidipine HCI resulted in distinct reduction of virus replication, the combined application of benidipine HCI and CQ further enhanced the anti-SARS-CoV-2 efficacy (P < 0.001).

**Figure 5.**
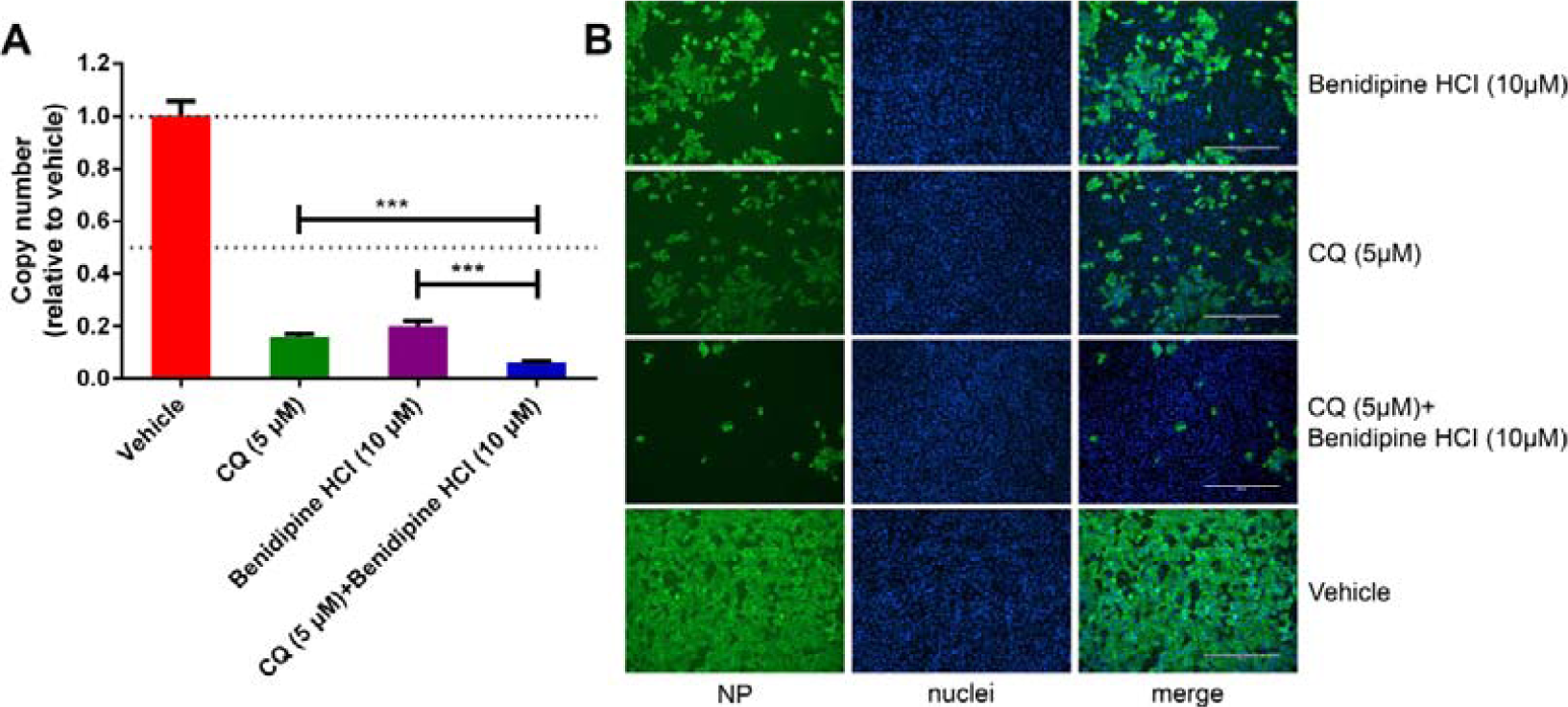
The antiviral activities of chloroquine (CQ) and/or benidipine HCI against SARS-CoV-2 replication. Vero E6 cells were treated with indicated concentrations of compounds separately or in combination and infected with SARS-CoV-2 at an MOI of 0.05. At 24 hours p.i., supernatant was collected and viral copy number in the supernatant was measured with quantitative RT-PCR (**A**), and NP expression in infected cells was analyzed by immunofluorescence with NP antibody (**B**). The experiments were performed in triplicates, and data shown are means ± SD. Comparison of mean values between two groups was analyzed by the student’s t test. *p < 0.05; **p < 0.01; ***p < 0.001. Bars: 400 μm.

### Administration of amlodipine besylate is associated with reduced case fatality rate in COVID-19 patients with hypertension

In order to evaluate whether CCBs have therapeutic effect in COVID-19 patients, we retrospectively analyzed the medical record of 487 adult COVID-19 patients with hypertension, including 225 had been admitted into the Tongji Hospital from January 17 to February 14,, and 262 had been admitted into the Union Hospital from January 10 to March 30, 2020. Of these patients 331 concurrently had other underlying comorbidities such as diabetes, chronic obstructive pulmonary disease, cerebral infarction, etc, 56 had no information on antihypertensive treatment, and 10 were still in the hospital. The 90 patients, who only had hypertension as the comorbidity and were either discharged from the hospital or deceased, were included for the retrospective analysis. Among these patients 44 received amlodipine besylate, 16 received nifedipine, 4 received other CCBs, 17 received other antihypertensive drugs (including ARBs, ACEIs, β-blockers, and thiazide), and 9 had no anti-hypertension drug treatment. No patient was found receiving benidipine HCI treatment. All the patients who did not receive amlodipine besylate were defined as non-amlodipine besylate treated patients. For amlodipine besylate treated and non-amlodipine besylate treated patients, the median (IQR) age was 67 (59.5-72) and 65 (57-74) years, and the median (IQR) delay from symptom onset to hospital admission was 10 (7-14) and 8.5 (6-13.5) days, respectively. Both of the two variables showed no significant inter-group difference. The female proportion was lower in amlodipine besylate treated patients (37.0%) than that (59.1%) in non-amlodipine besylat treated patients, (P = 0.036, Supplementary information, Table S1). The frequencies of clinical manifestations that were recorded before or at admission, including fever, cough, feeble, chest distress, shortness of breath, and gastrointestinal symptoms, were comparable between the two groups (all P > 0.05, Supplementary information, Table S1). Compared to the non-amlodipine besylate treated group, the amlodipine besylate treated group had lower serum levels of total bilirubin and lactate dehydrogenase (P = 0.047 and P = 0.015, respectively; Supplementary information, Table S1). All other laboratory parameters tested at admission were comparable. The commonly prescribed therapies during hospitalization included antibiotics, antiviral agents, traditional Chinese medicines, corticosteroids, and respiratory support. The amlodipine besylate treated group had lower frequency of antibiotics and higher frequency of corticosteroids (P = 0.028 and P = 0.006, respectively; Supplementary information, Table S2), while other therapies were observed with comparable frequencies between the two groups.

For the primary outcome of mortality, a beneficial effect in reducing the case fatality rate (CFR) was observed in patients receiving amlodipine besylate, with the CFR being significantly decreased from 26.1% (12/46) in non-amlodipine besylate treated group to 6.8% (3/44) in amlodipine besylate treated group (P = 0.022). Kaplan-Meier analysis similarly demonstrated reduced risk of death in amlodipine besylate treated group, in comparison with non-amlodipine besylate treated group (P = 0.033, log-rank test; Figure 6A). The effect amlodipine besylate treatment of on CFR remained significant with the use of Cox regression model by adjusting for age, sex, the delay from symptom onset to hospital admission, and therapies administration (hazard ratio (HR) 0.182, 95% confidence interval (CI) 0.037-0.897, P = 0.036; Table 1). Further analysis showed that the CFRs were higher in all other patient groups, with 12.5% (2/16) in patients receiving nifedipine, 25.0 (1/4) in patients receiving other CCBs, 41.2% (7/17) in patients receiving other anti-hypertension drugs, 22.2% (2/9) in patients without receiving anti-hypertension drugs. When compared to the patients without receiving anti-hypertension drugs, significant treatment effect was only observed in the patients receiving amlodipine besylate (Table 1; Figure 6B).

**Table 1.** Treatment effect of amlodipine besylate and other antihypertensive drugs in reducing mortality in the patients of COVID-19.

|  | Total<br>(n=90) | Survival<br>(n=75) | Fatal<br>(n=15) | *P*<sup>a</sup> | HR (95% CI) | *P*<sup>b</sup> | Adjusted<br>HR (95% CI) | *P*<sup>c</sup> |
| --- | --- | --- | --- | --- | --- | --- | --- | --- |
| Treatment regimen |  |  |  |  |  |  |  |  |
| No treatment | 9 | 7 (77.8) | 2 (22.2) | 0.026 | Reference |  | Reference |  |
| Amlodipine besylate | 44 | 41 (93.2) | 3 (6.8) |  | 0.141 (0.036-0.546) | 0.005 | 0.086 (0.014-0.551) | 0.010 |
| Nifedipine | 16 | 14 (87.5) | 2 (12.5) |  | 0.243 (0.050-1.174) | 0.078 | 0.213 (0.040-1.135) | 0.070 |
| Other CCBs | 4 | 3 (75.0) | 1 (25.0) |  | 0.483 (0.100-2.329) | 0.365 | 0.634 (0.104-3.853) | 0.621 |
| Other antihypertensive drugs | 17 | 10 (58.8) | 7 (41.2) |  | 0.470 (0.058-3.834) | 0.480 | 0.727 (0.078-6.792) | 0.780 |
| Amlodipine besylate |  |  |  |  |  |  |  |  |
| No | 44 | 41 (93.2) | 3 (6.8) | 0.022 | Reference |  | Reference |  |
| Yes | 46 | 34 (73.9) | 12 (26.1) |  | 0.253 (0.071-0.895) | 0.033 | 0.182 (0.037-0.897) | 0.036 |

Other antihypertensive drugs include angiotensin receptor blockers, angiotensin converting enzyme inhibitors, β-blockers, and thiazide.
<sup>a</sup>Analysed by Chi-square test or Fisher's exact test.
<sup>b</sup>Analysed by Kaplan-Meier model
<sup>c</sup>Analysed by Cox regression model by adjusting for age, sex, the delay from symptom onset to hospital admission, and therapies administration.

**Figure 6.**
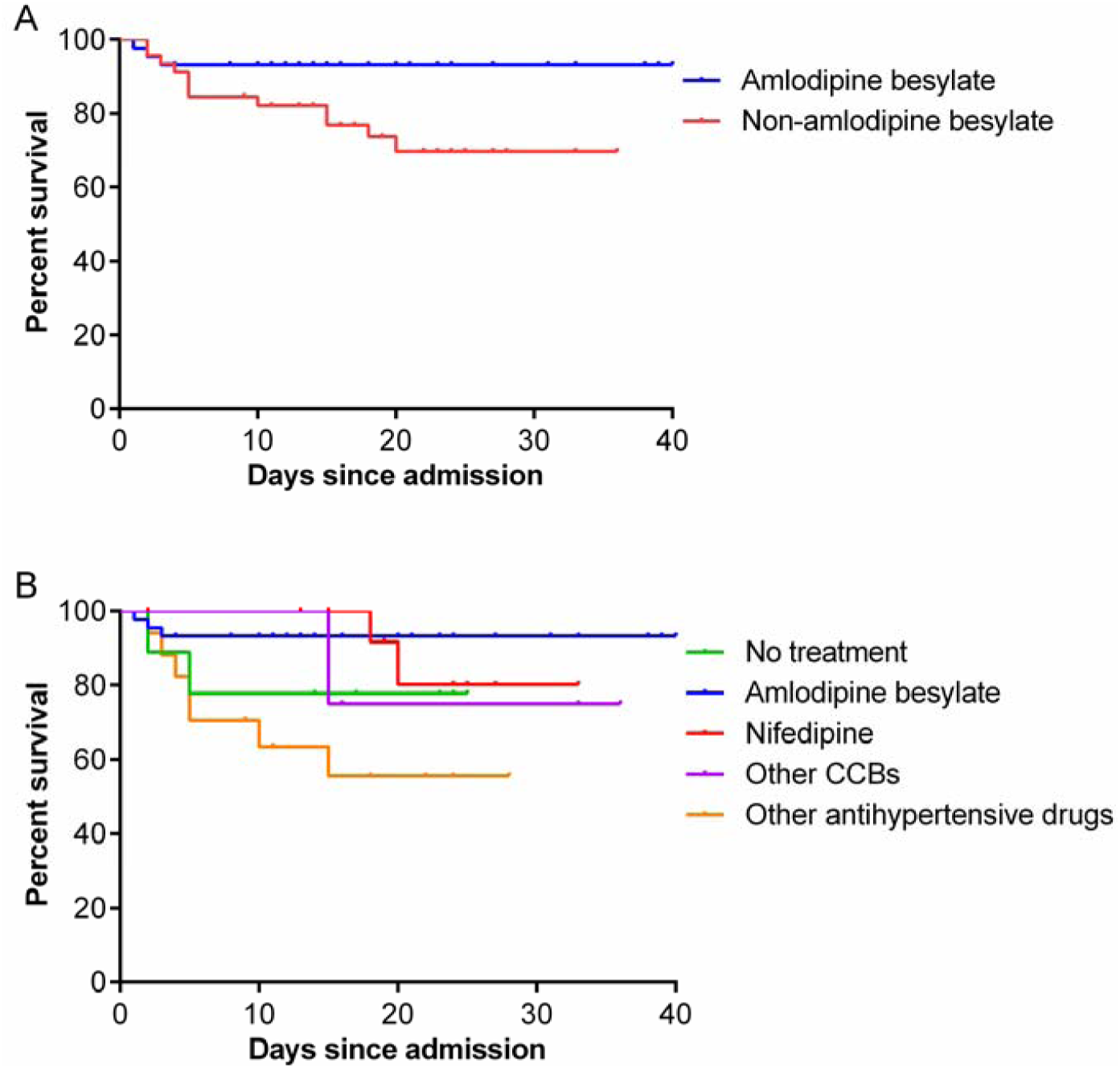
Analysis of amlodipine besylate treatment on probability of survival in COVID-19 patients with hypertension. Treatment effect on probability of survival of amlodipine besylate treated patients was compared with non-amlodipine besylate treated patients (**A**), or with patients received different types of anti-hypertension drugs (**B**). Other antihypertensive drugs include angiotensin receptor blockers, angiotensin converting enzyme inhibitors, β-blockers, and thiazide. The Kaplan-Meier method was used to analyze the time-to-event data.

## Discussion

Depending on the studies, around 13-30% of COVID-19 patients have hypertension as the underlying comorbidity (*11, 16, 19*). The case fatality rate of this group of patient is calculated to be 6%, which is more than 6-fold higher than the CFR of people without underlying comorbidity (0.9%) (*19*). In Wuhan, where the proportion of patients with critical conditions is higher, the CFR of patients with hypertension can be up to 14%. Effective medication is needed for treatment of this group of patients. ARBs, ACEIs and CCBs are three major types of anti-hypertension drugs that are widely used in the clinics. It was reported that the ARBs or ACEIs, such as losartan, olmesartan, lisinopril, etc, lead to significantly higher cardiac ACE2 mRNA level in animal model (*20, 21*). Since the SARS-CoV-2 virus uses ACE2 as its entry receptor, this raises the concern whether administration of these two types of drugs would lead to higher expression level of ACE2 and result in more severe virus infection (*22*). We showed here that, of the three types of anti-hypertension drugs, only CCBs such as, benidipine HCI or amlodipine besylate, showed potent anti-SARS-CoV-2 activity in vitro. The retrospective clinical investigation of 90 COVID-19 patients with hypertension further revealed the beneficial effect of amlodipine besylate administration with reduced CFR (6.8%, n=44). In contrast, patients received ARBs/ACEIs/-blockers/thiazide as anti-hypertension drugs had the CFR of 41.2% (n=17) and the general CFR of this group of patient is 16.7% (n=90). These results together suggest that CCBs, such as amlodipine besylate, may be more effective drug options for treating COVID-19 patients who have hypertension as the comorbidity.

The therapeutic mechanism of CCBs against COVID-19 still awaits further investigation. Several pathogenic viruses, such as Zika virus, dengue virus, H5N1 avian influenza virus, etc, induce intracellular calcium influx to facilitate virus infection (*23, 24*). The elevated intracellular calcium level is associated with pathogenesis mechanisms including induction of mitochondrial dysfunction and cell death which will result in triggering of strong inflammatory responses (*25-27*). Consistently, CCBs were reported to have anti-inflammatory efficacy through regulating intracellular calcium level in patients and to decrease mortality in septic animal models with excessive inflammatory responses (*28, 29*). Particularly, amlodipine besylate has been shown to decrease levels of inflammatory markers and oxidative stress compared to baseline in patients with hypertension (*30*). Excessive inflammatory responses are reported to be associated with COVID-19 fatal outcome (*11*). It is possible that, besides inhibiting virus replication, CCBs may also function through alleviating inflammatory responses in the patients to achieve the clinical benefits in a synergistic way with its anti-viral efficacy.

Recently, CCBs have been reported to inhibit replication of several emerging viruses including Ebola virus, Marburg virus (*31, 32*), Junin virus(*14*), and severe fever with thrombocytopenia syndrome virus (SFTSV) (*33*). Particularly, CCB treatment was reported to be associated with reduced CFR among SFTS patients (*13*). Here we show that, similar with SFTSV, CCBs inhibit the post-entry events of SARS-CoV-2 replication. Although the exact inhibition mechanism still needs further investigation, it is possible that CCBs block the virus-induced intracellular calcium influx and impair calcium dependent cellular pathways that are critical for virus replication. This way CCBs may function as a host-oriented drug that inhibits virus replication through regulating virus-dependent host machinery and the chance for occurrence of resistant mutants is lower compared to anti-viral drugs that target specific virus constituents (*34*). This would be highly valuable for developing drugs against RNA viruses such as SARS-CoV-2 as these viruses generally have a high mutation rate.

CQ has been shown to efficiently block SARS-CoV-2 entry in vitro and emerging evidences showed that administration of CQ has beneficial effects for COVID-19 patients in clinics. It was also reported that administration of CQ can reduce overall inflammation in several conditions with little toxicity (*9*). Whether CQ also alleviates the excessive inflammatory responses in COVID-19 patients is currently unknown. Nevertheless, the significantly enhanced anti-SARS-CoV-2 efficacy upon combined application of CQ and CCB indicates that dual administration of these two drugs may achieve a more pronounced therapeutic effect. Several clinical trials are currently ongoing for analyzing the therapeutic effect of CQ in COVID-19 patients. Whether there are patients that have received combined drug treatment of CQ and CCB would be interesting for evaluation.

Results from this study suggested that CCB amlodipine besylate is associated with reduced case fatality rate of COVID-19 patients with hypertension. COVID-19 patients with several comorbidities besides hypertension may have a more complicated underlying condition, and therefore was not included in the current study. Thus the therapeutic potential may only be applicable to the patients with hypertension as the only comorbidity. Evaluation with a larger patient cohort would further verify the potential therapeutic effect of the CCB. Additionally, dosing, side-effects and drug-drug interactions of the CCBs, similar with any drug that is in clinical use or testing, should be rigorously evaluated before clinical benefits can be more formally concluded.

## Materials and Methods

### Cells, virus and reagents

Vero E6 cell line was obtained from American Type Culture Collection (ATCC) and maintained in minimum Eagle’s medium (MEM; Gibco Invitrogen) supplemented with 10% fetal bovine serum (FBS; Gibco Invitrogen), 1% antibiotic/antimycotic (Gibco Invitrogen), at 37 °C in a humidified 5% CO_2_ incubator. Huh7 cell line was cultured in Dulbecco’s modified Eagle’s medium (DMEM; Gibco Invitrogen) supplemented with 10% FBS, 1% antibiotic/antimycotic (Gibco Invitrogen), at 37 °C in a humidified 5% CO_2_ incubator.

SARS-CoV-2 (nCoV-2019BetaCoV/Wuhan/WIV04/2019) was propagated in Vero E6 cells (*2*), and viral titer was determined by 50% tissue culture infective dose (TCID50) as described in our previous study(*5*). All the infection experiments were performed in a biosafety level-3 (BLS-3) laboratory.

Benidipine HCI (Selleck Chemicals, no. S2017), Amlodipine besylate (Selleck Chemicals, S1813), Cilnidipine (Selleck Chemicals, S1293), Nicardipine HCl (Selleck Chemicals, S4181), Nifedipine (Selleck Chemicals, S1808), Isradipine (Selleck Chemicals, S1662), Nimodipine (Selleck Chemicals, S1747), Nisoldipine (Selleck Chemicals, S1748), Felodipine (Selleck Chemicals, S1885), 2-Aminoethyl Diphenylborinate (2APB, Selleck Chemicals, S6657), BAPTA-AM (Selleck Chemicals, S7534) and Chloroquine (Sigma-Aldrich, no.C6628) were purchased from indicated companies.

### Evaluation of the antiviral activities of the test compounds

Vero E6 pre-seeded in 48-well dish (1 × 10^5^ cells/well) were treated with the different concentration of the indicated compounds for 1 hour and infected with SARS-CoV-2 at an MOI of 0.05. Two hours later, the virus-drug mixture was removed and cells were cultured with drug containing medium. At 24 hours p.i., the cell supernatant was collected and lysed. The viral RNA extraction and quantitative real time PCR (RT-PCR) analysis was described in our previous study (*5*).

### Evaluation of the cytotoxicity of the test compounds

Vero E6 pre-seeded in 96-well dish (5 × 10^4^ cells/well) were treated with the different concentration of the indicated compounds, and 24 hours later, the relative numbers of surviving cells were measured with cell counting kit-8 (GK10001,GLPBIO) according to the manufacturer’s instructions.

### Immunofluorescence microscopy

To detect intracellular expression level of viral NP, cells were fixed with 4% paraformaldehyde in advance. Fixed cells were permeabilized with 0.5% Triton X-100 and blocked with 5% bovine serum albumin (BSA).Then they were incubated for 2 hours with the anti-sera (1:1000 dilution) against the NP of a bat SARS-related CoV as the primary antibody, followed by incubation with Alexa 488-labeled goat anti-rabbit IgG (Abcam, ab150077; 1:500 dilution). The nuclei were stained with DAPI (Sigma-Aldrich, no.D9542). The images were taken by a fluorescence microscopy.

### Western blot analysis

For Western blot analysis, proteins were separated by 12% SDS-PAGE and then transferred onto PVDF membranes (Millipore). The membranes were blocked with 5% BSA in TBST (TBS buffer with 0.1% Tween 20) for 1 hour at room temperature. After washed with TBST for three times, the membranes were incubated with the anti-NP sera (1:2000 dilution) overnight at 4°C. After washed with TBST for three times, the membranes were incubated with horseradish peroxidase (HRP)-conjugated Goat Anti-Rabbit IgG (Proteintech, China; 1:10000 dilution). Protein bands were detected by SuperSignal West Pico Chemiluminescent substrate (Pierce).

### Clinical investigation

#### Study design and patients

To investigate the clinical effect of amlodipine treatment on COVID-19, we conducted a retrospective clinical investigation on the patients who were admitted to the Tongji Hospital, Union Hospital, which are the major tertiary teaching hospitals in Wuhan, China, and are responsible for the treatments of severe COVID-19 cases. The diagnosis of COVID-19 was made based on the World Health Organization interim guidance, and the confirmed cases denoted the patients whose nasal or pharyngeal swab samples were positive for real-time reverse-transcription polymerase-chain-reaction (RT-PCR) assay. Adult confirmed patients were checked for medical record of comorbidities and related therapeutic drugs by a trained research medical staff, and the COVID-19 patients who had hypertension were recruited into the study. Patients, who had other comorbidities, such as coronary heart diseases, cerebral infarction, diabetes, chronic obstructive pulmonary disease, pulmonary tuberculosis, chronic kidney disease, and malignancy, were excluded. The research protocol was approved by the human ethics committee of the hospital in accordance with the medical research regulations of China (TJ-IRB20200102), and oral informed consents were obtained from all patients or patients’ family members.

### Data collection

Data about demography, clinical manifestations, and laboratory testing results were retrospectively collected by reviewing medical records and entered into standardized database. Medication use during hospitalization including information on antihypertensive drugs (i.e. calcium channel blockers, angiotensin receptor blockers, and diuretics) was also recorded. Serial throat swabs were collected for the testing of HCoV-19 RNA with the use of RT-PCR during the patients’ hospitalization.

### Outcome

The primary outcome was case fatality.

### Statistics

Continuous variables were summarized as means and standard deviations or as medians and interquartile-range (IQR). Student’s *t* test or nonparametric test (Mann-Whitney test) was used as appropriate for comparisons of continuous variables between two groups, and ANOVA test or nonparametric test was used as appropriate for comparisons of continuous variables among multiple groups. Categorical variables were summarized as frequencies and proportions, and were analysed by Chi-square test or Fisher’s exact test as appropriate. We used the Kaplan-Meier method and the log-rank test to analyse time-to-event data for treatment effect analysis. We calculated HRs and 95% CI by using Cox regression models. A 2-sided *P* value of <0.05 was considered to be statistically significant. All statistical analyses were performed using SPSS software, version 19.0.

## Data Availability

All data in the study are available from the corresponding author by request

## Acknowledgements

We thank Hao Tang, Jia Wu, Jun Liu and Tao Du from BSL-3 Laboratory of Wuhan Institute of Virology for their critical support.

## Author contributions

L.-K.Z., K.P., and G.X. conceived and supervised the study. K.P., L.-K.Z., H.L., and G.X. wrote the manuscript. H.Z. collected clinical data. H.L., W.L., H.Z. and H.C. analyzed clinical data. Y.S., X.-M.J., W.-J.S., Y.W., S.L., and Y.-L.Z. performed in vitro experiment. L.-K.Z., K.P., Y.S., and H.L contributed to the design of the study and data analysis. All authors had access to the study data, and reviewed and approved the final manuscript.

